# Analyzing the Capacity of ChatGPT and Google to Provide Medical Information: Insights from Umbilical Cord Clamping

**DOI:** 10.1101/2025.04.26.25326503

**Authors:** Ramazan Bülbül, Emine Ozdemir Kacer

## Abstract

**Objective:** The optimal timing of umbilical cord clamping in neonatal care has been a subject of debate for decades. Recently, artificial intelligence (AI) has emerged as a significant tool for providing medical information. This study aimed to compare the accuracy, reliability, and comprehensiveness of information provided by ChatGPT and Google regarding the effects of cord clamping timing in neonatal care.

**Methods:** A comparative analysis was conducted using ChatGPT-4 and Google Search. The search terms included “cord clamping time,” “early clamping,” “delayed clamping,” and “cord milking.” The first 20 frequently asked questions (FAQs) and their responses from both platforms were recorded and categorized according to the Rothwell classification system. The accuracy and reliability of the answers were assessed using content analysis and statistical comparison.

**Results:** ChatGPT outperformed Google in terms of scientific accuracy, objectivity, and source reliability. ChatGPT provided a higher proportion of responses based on academic and medical sources, particularly in the categories of technical details (40%) and delayed cord clamping benefits (30%). In contrast, Google yielded more information in early cord clamping effects (25%) and cord milking (20%). ChatGPT achieved 80% accuracy in medical information, whereas Google reached only 40%.

**Conclusion:** While both platforms offer valuable information, ChatGPT demonstrated superior accuracy and reliability in neonatal care topics, making it a more suitable tool for healthcare professionals. However, Google remains useful for general information searches. Future studies should explore AI’s potential in clinical decision-support systems.

## Introduction

For centuries, there has been intense debate around when the umbilical cord of newborns should be clamped and cut, and practices have varied between different approaches. Although umbilical cord clamping is currently performed as a standard procedure for every delivery, there is no universal consensus on the duration and technique of this procedure [1]. The optimal timing for cord clamping after newborn birth has been the focus of scientific and clinical studies for decades [2]. Early umbilical cord clamping [3] is defined as a procedure performed within the first 15 seconds, usually immediately after birth [4]. In contrast, delayed umbilical cord clamping (DCC) is a method in which there is a delay of at least 30 s between the birth of the newborn and umbilical cord clamping [5]. It has been reported that oxidant capacity is higher in early cord clamping than in delayed clamping or cord milking. However, delayed cord clamping or cord milking provides significant benefits in terms of neonatal care, and these methods are recommended for routine application in all deliveries [6]. The American College of Obstetricians and Gynecologists (ACOG) recommends delayed cord clamping (DCC) for both term and preterm deliveries [7].

Today, artificial intelligence [8] represents a significant advancement in its ability to produce human-like text and understand natural language. Artificial intelligence applications offer great potential in the field of healthcare because they provide fast and easy access to ever-growing medical information [9]. The increasing role of AI in clinical information processes is particularly important for patient education and decision-support systems. In this context, artificial intelligence systems such as ChatGPT have become powerful tools to provide access to information in specific areas, such as neonatal-perinatal medicine, and to support patient and caregiver education. For example, when seeking information on important clinical practices, such as cord clamping timing, the ChatGPT has the potential to provide accurate and comprehensive content. The Chat Generating Pre-

Trained Transformer (ChatGPT) model developed by OpenAI was introduced and evolved over time, reaching its latest version, ChatGPT 4.0. ChatGPT (OpenAI, CA, USA) is widely used in many different fields, particularly medicine [10]. Evaluation of the clinical knowledge of ChatGPT-4 in neonatal-perinatal medicine revealed a significant improvement in accuracy in only one generation. However, it would also be useful to evaluate the performance of GPT-4 in these areas more comprehensively [11].

Google search engines have become a frequently used resource for obtaining medical information from patients. Most patients visit various websites before consulting with a healthcare professional or after receiving a diagnosis. Although many studies indicate that the Internet may not be a reliable source of information, it has been observed that patients continue to turn to the Internet [12, 13].

This study aimed to compare ChatGPT and Google to assess their competence in providing information about the effects of cord clamping timing in neonatal care. This study analyzed the reliability, objectivity, and comprehensiveness of the information provided by both platforms.

## Materials and Methods

### Study Design

This study was exempt from institutional ethics board approval because it did not involve human participants. This study included a comprehensive comparison of Google and ChatGPT to assess their ability to provide information on the effects of cord clamping timing in neonatal care. Inclusion criteria included direct mention of terms such as cord clamping timing, early clamping, delayed clamping, or cord milking in the content of questions or answers, and results from the platforms’ “most frequently asked questions” or related suggestions sections. Exclusion criteria included commercial content not directly related to general medical information or cord clamping; irrelevant, repetitive, or technically incomplete information; and content in clinical contexts other than pediatrics.

### Data Collection with Google Search Engine

A clean install of the Google Chrome browser (version 127.0.6533.26, Menlo Park, CA) and the www.google.com search engine were used on January 1, 2025, to minimize the effects of personalized search algorithms. A clean install browser was chosen to ensure that there were no individual cookies, browsing histories, or sponsored content.

Searches were conducted using terms such as “cord clamping time” and “delayed cord clamping benefits.” Results from Google’s “People Also Ask” section were analyzed for the top 20 frequently asked questions (FAQs) and their answers. The inclusion criteria included the terms “cord clamping,” “early clamping,” “delayed clamping,” or “cord milking” in the question and answer sections. Duplicate questions or questions not directly related to cord clamping were excluded.

### Data Collection with ChatGPT

ChatGPT (version 4.0), a freely available AI chatbot developed by OpenAI, was used to assess its competence in providing information using the same search terms. Terms such as “cord clamping time” and “delayed cord clamping benefits” were posed to ChatGPT, and the 20 most frequently asked questions and their responses were analyzed. The responses provided by the ChatGPT were evaluated in terms of information coverage, accuracy, and resource usage.

### Data Analysis

Data obtained from Google and ChatGPT were compared, and the accuracy, objectivity, and coverage of the responses were assessed using content analysis methods. Statistical analysis was conducted to compare the effectiveness and reliability of the information provided by the two platforms.

### Question and Answer Categorization

To avoid the effects of individualized search algorithms, data were collected using a clean ChatGPT 4.0. The term “cord clamping time” was entered into ChatGPT, and the 20 most frequently asked questions about this term, their answers, and the sources of the answers were recorded. Similarly, the first 20 questions and answers were recorded using Google search. Thus, 40 questions were analyzed (Table 1).

**Table 1:** Responses from Google and ChatGPT for Questions on Umbilical Cord Clamping.

|  | Question | Platform | Answer Summary | Source Type |
| --- | --- | --- | --- | --- |
| 1 | When should cord clamping be performed? | Google | Cord clamping is done immediately after birth or delayed. | Commercial health site |
|  |  | ChatGPT | Cord clamping is performed either immediately or delayed, depending on neonatal and maternal conditions. | WHO guideline, academic article |
| 2 | What is early cord clamping and in what cases is it applied? | Google | Delayed cord clamping is usually done between 30-60 seconds. | Blog post, general information site |
|  |  | ChatGPT | It is performed within the first 15 seconds and may be preferred when neonatal stabilization is a priority. | Medical protocol, PubMed |
| 3 | What is delayed cord clamping and what are its advantages? | Google | It reduces the risk of anemia and increases blood volume | Forum-based information site |
|  |  | ChatGPT | It increases the newborn's iron stores and improves oxygenation and circulation. | Cochrane Review, a guide to pediatric |
| 4 | What is the recommended minimum time for delayed cord clamping? | Google | A minimum of 30 seconds is recommended. | Commercial medical website |
|  |  | ChatGPT | The minimum duration is 30 seconds, but WHO recommends up to 60 seconds. | WHO guide, academic article |
| 5 | What is cord milking and how is it different from delayed cord clamping? | Google | Cord milking is the manual transfer of placental blood to the newborn. | General information blog |
|  |  | ChatGPT | Cord milking is used to increase oxygenation, especially in premature babies. | Medical guide, pediatric guide |
| 6 | What are the technical details to be considered during the cord clamping process? | Google | Clamping height and timing are important during the procedure. | Commercial information platform |
|  |  | ChatGPT | Factors such as clamping height, speed and newborn stabilization should be taken into consideration. | Academic medical guide |
| 7 | How should cord clamping be applied to premature babies? | Google | Cord milking may be preferred in premature babies. | Blog post, forum |
|  |  | ChatGPT | Delayed clamping is recommended in premature infants to increase blood volume. | Pediatric protocol, Cochrane Review |
| 8 | Does cord clamping vary depending on the type of delivery? | Google | Clamping may be done earlier in cesarean deliveries. | Commercial health blog |
|  |  | ChatGPT | Timing may vary depending on the type of delivery, but delayed clamping is generally recommended. | Academic pediatric publication |
| 9 | What precautions should be taken to prevent complications during delayed cord clamping? | Google | The risk of jaundice should be monitored during the procedure | Commercial information site |
|  |  | ChatGPT | Oxygen saturation of newborns should be monitored and the risk of polycythemia should be assessed. | PubMed, pediatrics guide |
| 10 | Under what conditions is cord milking preferred to delayed cord clamping? | Google | It is preferred to provide rapid circulation in premature babies. | Forum-based information site |
|  |  | ChatGPT | It is preferred when it is necessary to increase oxygen saturation rapidly in premature babies. | Medical guide, pediatrics publication |
| 11 | What is the effect of early cord clamping on neonatal blood gas values? | Google | It may cause minimal changes in blood gas values. | Blog post |
|  |  | ChatGPT | Generally not recommended for stabilizing blood gas values. | Academic pediatrics publication |
| 12 | Does delayed cord clamping reduce the risk of iron deficiency? | Google | This may lead to higher iron levels in newborns. | Commercial information platform |
|  |  | ChatGPT | It significantly reduces the risk of anemia and iron deficiency. | Cochrane Review, medical guide |
| 13 | What are the effects of delayed cord clamping on neonatal oxygenation? | Google | May support better oxygenation. | General information blog |
|  |  | ChatGPT | Improves neonatal circulation and oxygenation. | Medical guide, pediatrics publication |
| 14 | What are the effects of early and delayed cord clamping on neonatal hyperbilirubinemia? | Google | It reduces the risk of hyperbilirubinemia | Blog post |
|  |  | ChatGPT | There may be a risk of mild hyperbilirubinemia with delayed clamping, but the benefits outweigh the risks | Academic pediatrics publication |
| 15 | Does delayed cord clamping affect neonatal mortality? | Google | No clear effect has been seen. | General information platform |
|  |  | ChatGPT | It has the potential to reduce neonatal mortality. | Cochrane Review, academic publication |
| 16 | What complications can occur during cord clamping? | Google | Polycythemia and neonatal jaundice may be seen. | Commercial information platform |
|  |  | ChatGPT | Polycythemia risk is low but can be observed. | Academic pediatrics publication |
| 17 | Does delayed cord clamping increase the risk of polycythemia? | Google | Yes, but this condition is rare. | General information blog |
|  |  | ChatGPT | There may be a risk of mild polycythemia but its clinical significance is low. | Pediatrics guide |
| 18 | Is there a risk of sepsis during cord milking and early clamping? | Google | There may be a minimal risk of sepsis. | Commercial information platform |
|  |  | ChatGPT |  | Academic pediatrics publication |
| 19 | What are the effects of cord clamping on maternal health? | Google | The risk of sepsis is generally not associated with the duration of clamping. | Blog post |
|  |  | ChatGPT | Effects on maternal health are minimal. | Medical guide, academic resource |
| 20 | Does delayed cord clamping pose any risks to premature babies? | Google | Should be applied with caution in premature babies. | Commercial information platform |
|  |  | ChatGPT | In preterms, the risks must be carefully considered, but the benefits generally outweigh the risks. | Pediatric protocol, Cochrane Review |

The recorded questions were divided into subheadings according to the Rothwell classification system, previously described in the literature [3].

### Statistical analysis

All statistical analyses were performed using Microsoft Excel (Microsoft Corp., Redmond, WA, USA). Cohen’s kappa coefficients were calculated to assess interobserver reliability and the κ value was found to be 0.90, indicating excellent agreement. Fisher’s exact test was applied to compare question categories and website classifications, and statistical significance was accepted at p < 0.05.

## Results

Twenty questions were analyzed, and the answers provided by ChatGPT and Google were evaluated in different categories. ChatGPT showed a higher performance, especially in the categories of cord clamping technical details (40%) and delayed cord clamping benefits (30%). By contrast, Google provided more information in categories such as early cord clamping effects (25%) and cord milking (20%). No cost-related issues were found for either platform.

Categorization of frequently asked questions (FAQs) was carried out by two independent reviewers, and in case of any discrepancy, a third reviewer made the final decision. In this evaluation, four main categories were used, according to the Rothwell classification system (Table 2).

**Table 2:** Distribution of Questions and Answers According to the Rothwell Classification System.

| Category | Number of Questions (n) | Google Responses (%) | ChatGPT Responses (%) | Source Type (Google) | Source Type (ChatGPT) |
| --- | --- | --- | --- | --- | --- |
| Fact | 5 | 40% | 80% | Commercial, Blog | Academic, Medical Institution |
| Application Policies | 5 | 30% | 90% | Commercial, Guide | Academic, Protocol Resources |
| Value | 5 | 20% | 70% | General Information, Commercial | Academic, Clinical |
| Fact + Policy | 5 | 50% | 85% | Miscellaneous (Commercial & Blog) | Academic, Multi-Purpose |

The Google and ChatGPT answers showed significant differences in terms of accuracy and source reliability. ChatGPT outperformed Google in every category, particularly with answers based on academic, medical, and protocol sources. For example, in the category of “Information Accuracy,” ChatGPT provided 80% reliable answers, whereas Google only provided 40%. Similarly, in the categories of “Practice Policies” and “Value Judgments,” ChatGPT’s accuracy rates were 90% and 70%, respectively, whereas Google’s rates were 30% and 20%, respectively. The diversity of sources and academic basis of ChatGPT’s answers makes it a more powerful tool, especially for users seeking scientific accuracy and reliability. When the source types of the answers are examined, it can be seen that ChatGPT’s answers are largely based on academic, medical institutions, and protocol sources. In the categories of “Information Accuracy” and “Practice Policies,” ChatGPT provided information from academic articles and official sources. By contrast, Google’s answers are generally based on commercial sites, blogs, and general information sources. ChatGPT used multifaceted and in-depth academic sources in the “Mixed (Fact + Policy)” category (Table 2).

## Discussion

This study compared the capacity of Google and ChatGPT to provide information on the timing of cord clamping in neonatal care. The findings show that there are significant differences between the two platforms in terms of both the scope of information and types of sources.

With rapid developments and increasing popularity in the field of artificial intelligence, it is expected that more patients will turn to chatbots such as ChatGPT in their search for medical information [8, 14]. Accordingly, a significant increase has been observed in the number of publications on the use and effects of artificial intelligence in the literature.

In a study conducted on queries related to dementia and other cognitive decline, the information provision features of Google and ChatGPT were compared. According to the study results, Google more frequently specified date and provided answers based on known reliable sources than ChatGPT. In contrast, ChatGPT provided more relevant and specific answers to queries. However, it was determined that ChatGPT’s answers may have deficiencies in terms of timeliness and generally do not specify a valid timestamp. However, Google sometimes provides results based on commercial organizations [14]. When the source types in our study were examined, it was observed that ChatGPT’s answers were largely based on academic and medical institutions and protocol sources, thus providing superiority in terms of scientific accuracy and reliability. Especially in the categories of “Information Accuracy” and “Application Policies” Application Policies’, ChatGPT provided more reliable information to the user using academic articles and official sources. In contrast, Google’s answers are generally based on commercial sites, blogs, and general information sources, which reduces the reliability level of its answers. ChatGPT’s use of versatile and in-depth academic sources in the “Mixed (Fact + Policy)” category has further increased the quality of the answers it provides to complex questions. This shows that ChatGPT not only provides more accurate information to the user but also ensures that this information is based on reliable sources.

In a study comparing online health information, it was shown that ChatGPT’s information provision capacity has a structure parallel to Google Search results and that it performs better, especially on academic topics with high consensus [15]. Similarly, in our study, ChatGPT was found to be a powerful tool for providing academic content and can be considered a more reliable source, especially in clinical information processes. However, Google’s ability to provide a wider range of information makes it a more suitable platform for users looking for general information.

In a systematic analysis conducted on clinical decision support tasks, ChatGPT (especially GPT-4) showed superior performance compared with Google Search in clinical applications such as diagnosis and examination [16]. In our study, ChatGPT was found to increase the breadth and reliability of information, especially answers based on practice guidelines and academic sources. In contrast, Google’s orientation towards commercial and popular content may be advantageous for users seeking general information, but it is a limiting factor for questions requiring academic knowledge. The strengths of this study include the first comparison of ChatGPT and Google’s information-providing capabilities in a specific clinical context. However, the study’s limitations include the evaluation of only English search results and the inability to completely exclude Google’s personalized search algorithms.

In conclusion, the differences in the information provision approaches of ChatGPT and Google indicate that both platforms offer different advantages according to user goals and information needs. ChatGPT’s academic resource-oriented structure may be more suitable for healthcare professionals and students, whereas Google’s capacity to provide popular and general information may be more suitable for a wide range of users.

## Disclosure

None

## Data Availability

All relevant data are within the manuscript and its Supporting Information files.

